# Reliability and validity of the Japanese version of the Mental Health Literacy Scale (MHLS) among undergraduate and graduate students in the medical field

**DOI:** 10.1101/2022.02.01.22269195

**Authors:** Moeka Ikeyama, Kotaro Imamura, Norito Kawakami

## Abstract

**Objective:** Mental health literacy (MHL) is a fundamental basis of reducing stigma towards mental disorders and promoting early help-seeking. However, there is no internationally standardized scale covering all the attributes of MHL available in Japan. This study aimed to examine the reliability and validity of the Japanese version of the Mental Health Literacy Scale (MHLS) developed by O’Connor et al. (2015).

**Methods:** The MHLS was translated in accordance with international guidelines. Japanese students in the medical field were invited to complete an online questionnaire twice at baseline and two-week follow-up. Using the data, Cronbach’s alphas, intra-class correlation coefficient (ICC), and measurement errors were calculated for internal consistency and test-retest reliability. Correlations with mental health-related behavior (RIBS), accurate knowledge about mental disorders (MIDUS), intentions to seek help (GHSQ), and psychological distress (K6) were used to examine the convergent validity. Confirmatory factor analysis (CFA) and exploratory factor analysis (EFA) were performed to test structural validity.

**Results:** A total of 183 and 150 students responded at baseline and follow-up. Cronbach’s alpha coefficient was 0.76 and the ICC was 0.77 for the total score. The MHLS had a strong positive correlation with RIBS and moderate positive correlations with MIDUS and GHSQ. The CFA did not show a good fit either for one-or six-factor models, and the EFA yielded a four-factor structure.

**Conclusions:** The Japanese version of the MHLS demonstrated adequate reliability and validity, while the factor structure did not fit previously proposed models. This scale may be useful for assessing the degree of MHL among medical field students in Japan.

## Introduction

According to the World Mental Health Surveys, the quartile range of lifetime prevalence of mental disorders in the world is 18.1% to 36.1%^1)^, and the lifetime prevalence in Japan is reported to be 22.3%^2)^. Mental disorders are widespread worldwide, with detrimental personal, social, and economic consequences^3)^. For example, it has been reported that the percentage of people with mental disorders among the homeless is over 50%. Mental disorders have also resulted in the highest unemployment rate of all disabilities (up to 90%) and have been found to lead to poverty for individuals and families^4)^. However, 76% to 85% of people with severe mental disorders in low- and middle-income countries, and 35% to 50% in high-income countries, do not receive treatment^4)^. A study examining barriers to seeking help has shown that stigma is the most important barrier^5)^. The prejudice and discrimination against people with mental disorders are still an issue in the international community, and efforts to eliminate stigma are ongoing in Japan^6)^. As with other diseases, it is reported that early and appropriate treatment of mental disorders is effective for life prognosis^7,8)^, so reducing stigma and promoting early medical attention is necessary.

One of the most important concepts to reduce stigma and promote early reception is mental health literacy (MHL)^9,10)^. MHL refers to knowledge and attitudes about mental health that help people recognize, manage, and prevent mental health problems^11)^. According to Jorm et al.^11)^, MHL consists of six attributes: a) the ability to recognize specific disorders or different types of psychological distress; b) knowledge and beliefs about risk factors and causes of mental illness; c) knowledge and beliefs about self-help interventions; d) knowledge and beliefs about professional help available; e) attitudes which facilitate recognition and appropriate help-seeking; and f) knowledge of how to seek mental health information. The importance of MHL has been asserted because people with higher MHL have been found to have less prejudice, more willingness to interact with people with mental disorders^12)^, and higher intentions to seek help from professionals^13)^.

The scales to measure MHL developed so far can be broadly divided into vignette interviews and scale-based measures. Vignettes have some limitations. First, it is hard to understand a person’s MHL levels intuitively and concretely and to compare them between individuals. Second, they may answer the question based on a false identification because participants need to identify the mental disorders depicted in the vignette before answering the question. Third, this method requires time and effort because it involves interviews^14)^. Another method of measuring MHL, scale-based measurement, has also been developed in a variety of ways. For example, the Multiple-Choice Mental Illness Knowledge Test (MC-KOMIT) developed by Compton et al. in 2012^15)^, and the Mental Health Knowledge Schedule developed by Evans-Lacko et al. in 2010^16)^. These scale-based instruments are superior to vignettes in that they are more appropriate, more time-efficient, and provide more objective results. However, most scales do not measure all the six components of MHL^14)^. In 2015, O’Connor et al.^17)^ developed the Mental Health Literacy Scale (MHLS) in Australia. The MHLS consists of 35 items covering all attributes of MHL with univariate structure and is a methodologically and psychometrically superior measurement. Internal consistency, test-retest reliability, and construct validity have been demonstrated. Since the MHLS was developed in 2015, it has been used to measure MHL in various countries, and the MHLS has also been reported to be positively correlated with help-seeking (General Help-Seeking Questionnaire; GHSQ)^18)^ and quality of life (SF-12)^19)^. Furthermore, the MHLS has been translated into Vietnamese^20)^, Persian^21)^, Arabic^22)^, and Chinese^23)^. These studies indicate that the MHLS is reliable, valid, and applicable to a variety of populations. However, a Japanese version of the MHLS has not yet been developed.

The MHL of students in the medical field is important for several reasons. The first is that the stress level of medical students is high. It has been reported that the stress level of nursing students increases during their school years^24)^ and that medical students have higher psychological stress than their generations^25,26)^. However, it has been shown that many students feel the need to hide their mental health issues, which may be a barrier to seeking help^27)^. Increasing MHL may help them become more aware of their own problems and more likely to seek help, which may lead to better mental health. The second is that they will have many opportunities to come in contact with patients with mental disorders as medical professionals. It has been found that about 50% of people being treated for anxiety and depressive disorders were treated in general medical institutions^28)^ and that about 70% of people who die by suicide had contact with a general doctor in the last month^29)^. However, it has also been reported that mental disorders often go undiagnosed in primary care settings^28)^ and that over 85% of cases of mental disorders are missed by general physicians^30)^. From the above, it can be said that improving the MHL of medical students is important not only to improve their health problems but also to improve the quality of medical care with correct knowledge and responses.

This study aimed to investigate the reliability and validity of the newly developed Japanese version of the MHLS among Japanese students in the medical field. The internal consistency, test-retest reliability, structural validity, and convergent validity of the Japanese version of the MHLS were tested. We hypothesized that the Japanese version of the MHLS would have good internal consistency and test-retest reliability. Based on correlations for the original MHLS, we hypothesized that MHL scores measured by the MHLS would have weak-to-moderate correlations with intentions to seek help and no correlation with psychological distress. We also hypothesized that MHL scores would positively correlate with mental health-related behavior and accurate knowledge about mental disorders. Since a one-factor model was proposed in the original paper and the MHLS was created to include the six attributes of MHL, we assumed that a one-factor model or a six-factor model was appropriate. This paper was based on the COnsensus-based Standards for the selection of health Measurement Instruments (COSMIN) reporting guidelines^31)^.

## Methods

### Development of the Japanese version of the MHLS

The Japanese version of the MHLS was developed according to the procedure specified in the International Society of Pharmacoeconomics and Outcomes Research (ISPOR) task force guidelines^32)^ in the following five steps. (1) Preparation: we obtained permission from the original MHLS developers to translate into Japanese. (2) Forward-translation and reconciliation: two independent researchers, 1st author and a Japanese student in the medical field, translated from English into Japanese, and two translated sentences were compared and merged into one translated sentence. (3) Back-translation: an English speaker who did not know the original version of MHLS translated the Japanese version into English. (4) Back-translation review and harmonization: the original developer checked the back-translated measure and made revisions to ensure that the translation has the same meaning as the original version. (5) Cognitive debriefing: five

Japanese students were asked to complete the harmonized measure. Their feedback about the ease of answering, usability, and difficulties in understanding the measure was used for further revision. The results of these steps were integrated to create the final scale. The full version of the MHLS Japanese version can be found at (https://plaza.umin.ac.jp/heart/).

### Data collection

Online questionnaire surveys were conducted at baseline (August-October 2021) and two-week follow-up (August-November 2021) of students in the medical field in Japan. The study protocol was approved by the research ethics committee of the Graduate School of Medicine and the Faculty of Medicine, The University of Tokyo, Japan (No. 2021112NI).

### Participants

The participants were recruited through a snowball process using social media (email, line). Participant inclusion criteria were (a) Japanese or native Japanese speakers and (b) undergraduate and graduate students in the medical field. The exclusion criteria were those who were involved in the development process of the Japanese version of the MHLS. Based on these criteria, we asked students belonging to the target universities and graduate schools (around fifteen) to cooperate in the survey through our acquaintances. Participants were allowed to answer the self-report questionnaire by checking the consent box at the beginning of the questionnaire. Informed consent was taken through the questionnaire instructions, which explained that the protection of personal information was guaranteed and that personally identifiable information would be removed from the data.

### Measurements

Participants responded to an online self-report survey that included questions about MHL, mental health-related behaviors, accurate knowledge of the mental disorders, intention to seek help, psychological distress, and demographic variables.

#### Mental Health Literacy

The MHLS consists of 35 items, including all the attributes of MHL. 15 items were rated on a 4-point scale, and the other 20 items were rated on a 5-point scale^17)^. The total score (35 to 160) was calculated, and the higher the score indicates the higher MHL.

#### Mental health-related behavior

Mental health-related behavior was measured by the Japanese version of the Reported and Intended Behaviour Scale (RIBS)^33)^. The RIBS is an eight-item scale consisting of two subscales: (1) four items related to past or present contact with people with mental health problems and (2) four items related to participants’ future behavioral intentions when in contact with someone with mental health problems. The first subscale is scored as 1 = yes and 0 = no/don’t know (subscale score range: 0-4), with higher scores indicating more past or present contact. The second subscale is scored as 5 = agree strongly, 4 = agree slightly, 3 = neither agree nor disagree/don’t know, 2 = somewhat disagree, and 1 = strongly disagree (subscale score range: 4-20), with higher scores indicating more favorable behavioral intentions. The reliability and validity of the

Japanese version of the RIBS were confirmed in a previous study^33)^.

#### Accurate knowledge about mental disorders

The Mental Illness and Disorder Understanding Scale (MIDUS) assessed accurate knowledge about mental disorders^34)^. The MIDUS consists of 15 items and three subscales (treatability of illness, efficacy of medication, and social recognition of illness). All items are rated on a 5-point Likert scale (0 = Strongly agree, 4 = Strongly disagree). Lower total scores indicate better understanding. The reliability and validity of the MIDUS were confirmed in a previous study^34)^.

#### Intentions to seek help

Intentions to seek help were measured by the General Help-Seeking Questionnaire (GHSQ)^35)^. The GHSQ is a scale that measures intentions to seek help from various sources. The question was, “Imagine you are experiencing a personal-emotional problem or mental health difficulty”. For each of the sources presented, responses were made using the 7-point Likert scale (1 = extremely unlikely, 7 = extremely likely). Higher score indicates greater intentions to seek help.

#### Psychological distress

The Japanese version of the Kessler’s Psychological Distress Scale (K6) was used to assess psychological distress^36)^. This scale consists of six items that ask how often participants have experienced symptoms of psychological distress in the past 30 days. All items were scored on a 5-point Likert scale (0 = None of the time, 4 = All the time). The reliability and validity of the K6 were confirmed in a previous study^36)^.

#### Demographic variables

Demographic variables included gender, age, and educational background, whether undergraduate or graduate.

### Analysis

Several statistical values (Cronbach’s alphas, Intraclass Correlation Coefficients, the Standard Error of Measurement, and the Smallest Detectable Change) were used to test reliability. To test structural validity, the confirmatory factor analysis (CFA) and the exploratory factor analysis (EFA) were performed. To test convergent validity, correlational analyses were conducted. R version 4.0.1 (R Core Team, 2020) was used for each analysis.

To assess internal consistency, Cronbach’s alphas were calculated for the total score of the Japanese MHLS. Based on previous research, the sample size of more than 100 was considered sufficient for methodological quality for Cronbach’s alpha.

The Intra-class Correlation Coefficient (ICC) for the total score was calculated to assess test-retest reliability across the two weeks period. In addition, as the standards of measurement error, the Standard Error of Measurement (SEM) and the Smallest Detectable Change (SDC) were calculated. The SEM represents the standard deviation of repeated measures in a single participant and was calculated by computing the square root of the participant’s within-subject variance (SEM = √σbetween measurement + σresidual)^37)^. The SDC represents the smallest change that one participant must show in a measurement to ensure that the observed change is a true change and not just a measurement error and was calculated as 1.96 × √ (2 × SEM)^38)^.

The Kaiser-Meyer-Olkin (KMO)^39)^ measure of sampling fit and Bartlett’s test of sphericity were used to examine whether the data were suitable for factor analysis. If the KMO measure met the criterion (0.6 or higher) and the Bartlett’s test of sphericity had a p-value of less than 0.05, factor analysis was considered appropriate. Based on a previous study^40)^, factor analysis also requires a sample size of at least five to seven times the number of items and more than 100. Considering that the Japanese version of the MHLS has 35 items, the number of subjects in this study should be 175 or more.

The CFA was conducted to validate the one-factor and six-factor structural validity because it was reported in the original paper^17)^ that the MHLS assumed a univariate structure and that it was created to include the six attributes of MHL. The following indicators were used to evaluate the model fit: the chi-square (χ^2^), the Comparative Fit Index (CFI), the Tucker-Lewis Index (TLI), the Root Mean Square Error of Approximation (RMSEA), and the Standardized Root Mean Square Residual (SRMR). The goodness of fit was assessed using a combination of goodness-of-fit indicators, which included the ratio of χ2 to df (≤ 2), CFI > 0.90, TLI > 0.90, RMSEA < 0.08, and SRMR < 0.08. If the fit of the CFA outcome was not satisfactory, the EFA was performed using the generalized least-squares method and Oblimin rotation. The minimum average partial correlation (MAP) method was used to determine the number of factors.

To examine convergent validity, Pearson’s correlation coefficients (r) among the MHLS, the RIBS, the MIDUS, the GHSQ, and the K6 were calculated.

## Results

### Characteristics of participants

A flow chart of the participants is shown in **Fig 1**. The baseline response rate could not be determined because the number of people we could approach could not be ascertained. In the follow-up survey, 150 of 183 students responded (response rate = 82.0%). All items were set as required response items in the Internet-based survey, so there were no missing values for any variable or item. The demographic characteristics of the participants at baseline are shown in **Table 1**. In the baseline survey, there were more female than male respondents. The age range was 19-51 years (mean = 24.3, standard deviation [SD] = 5.5).

**Table 1.** Demographic characteristics of the participants (N = 183)

|  | Baseline survey<br>(N = 183) |  |
| --- | --- | --- |
|  | n (%) | Mean (SD) |
| Gender |  |  |
| Men | 76 (41.5%) |  |
| Women | 104 (56.8%) |  |
| Do not answer | 3 (1.6%) |  |
| Age, years |  | 24.3 (5.5) |
| Education |  |  |
| Undergraduate | 121 (66.1%) |  |
| Graduate | 62 (33.9%) |  |
| Mental health-related behavior (RIBS) |  | 16.3 (3.1) |
| Past or present contact with people with mental health problems |  | 1.1 (1.1) |
| Future behavioral intentions when in contact with someone with mental health problems |  | 15.2 (2.7) |
| Accurate knowledge about mental disorders (MIDUS) |  | 16.2 (7.2) |
| Treatability of illness |  | 3.4 (2.9) |
| Efficacy of medication |  | 10.9 (4.2) |
| Social recognition of illness |  | 2.0 (2.1) |
| Intentions to seek help (GHSQ) |  | 31.0 (7.6) |
| Psychological distress (K6) |  | 4.8 (4.7) |

RIBS: Reported and Intended Behaviour Scale, MIDUS: Mental Illness and Disorder Understanding Scale, GHSQ: General Help-Seeking Questionnaire.

**Fig 1.**
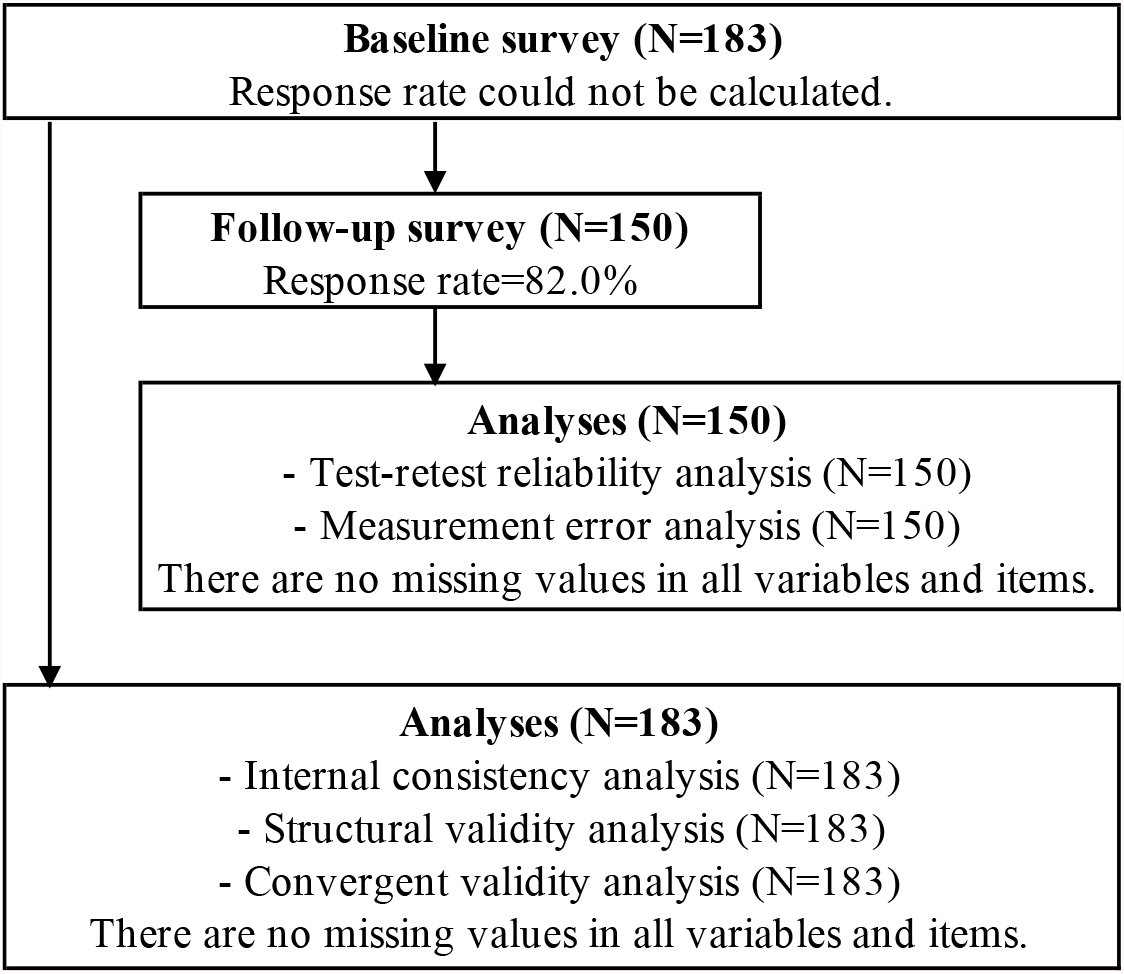
Flow chart of the participants

### Internal consistency and test-retest reliability

**Table 2** shows mean scores, Cronbach’s alphas (α), ICC, SEM, and SDC for the Japanese version of the MHLS. Cronbach’s alpha coefficient was 0.76. The ICC was 0.77, which means that about 80% of the variance of the two measurements was individually explained. SEM and SDC were 4.43 and 5.84, respectively.

**Table 2.**
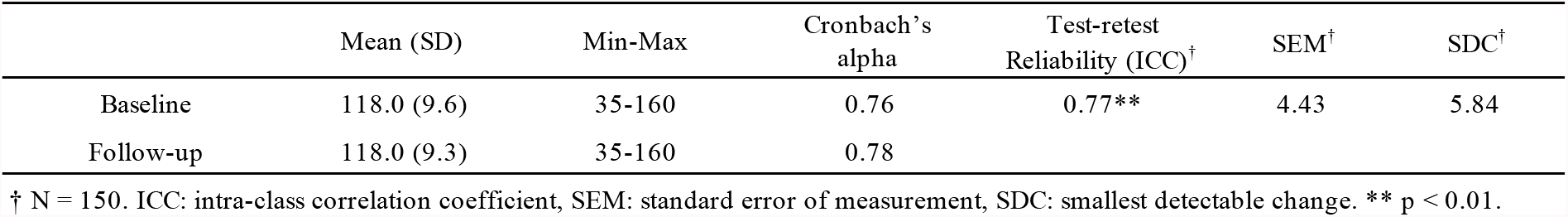
Mean scores, internal consistency, and reliability of the Japanese version of the MHLS (N = 183)

### Structural validity

The KMO score was 0.70, and Bartlett’s test of sphericity was significant (χ2 = 1874.823, df = 595, p < 0.01), indicating that factor analysis was suitable. The sample size of this study (N = 183) also met the criteria. The results of the CFA are shown in **Table 3**. Neither the one-factor model nor the six-factor model was a good fit. To investigate the factor structure, the EFA was conducted on 35 items. The MAP method indicated the four-factor structure. The generalized least-squares method with Oblimin rotation was used for the factor extraction, and the results are shown in **Table 4**. Given the six attributes of MHL, the three attributes (b: knowledge of risk factors and causes, c: knowledge of self-treatment, d: knowledge of professional help available) were not combined into one factor. The items of knowledge of how to seek information (f) belonged to one factor, and the items of attitudes that promote recognition or appropriate help-seeking behavior (e) were divided into two factors.

**Table 3.**
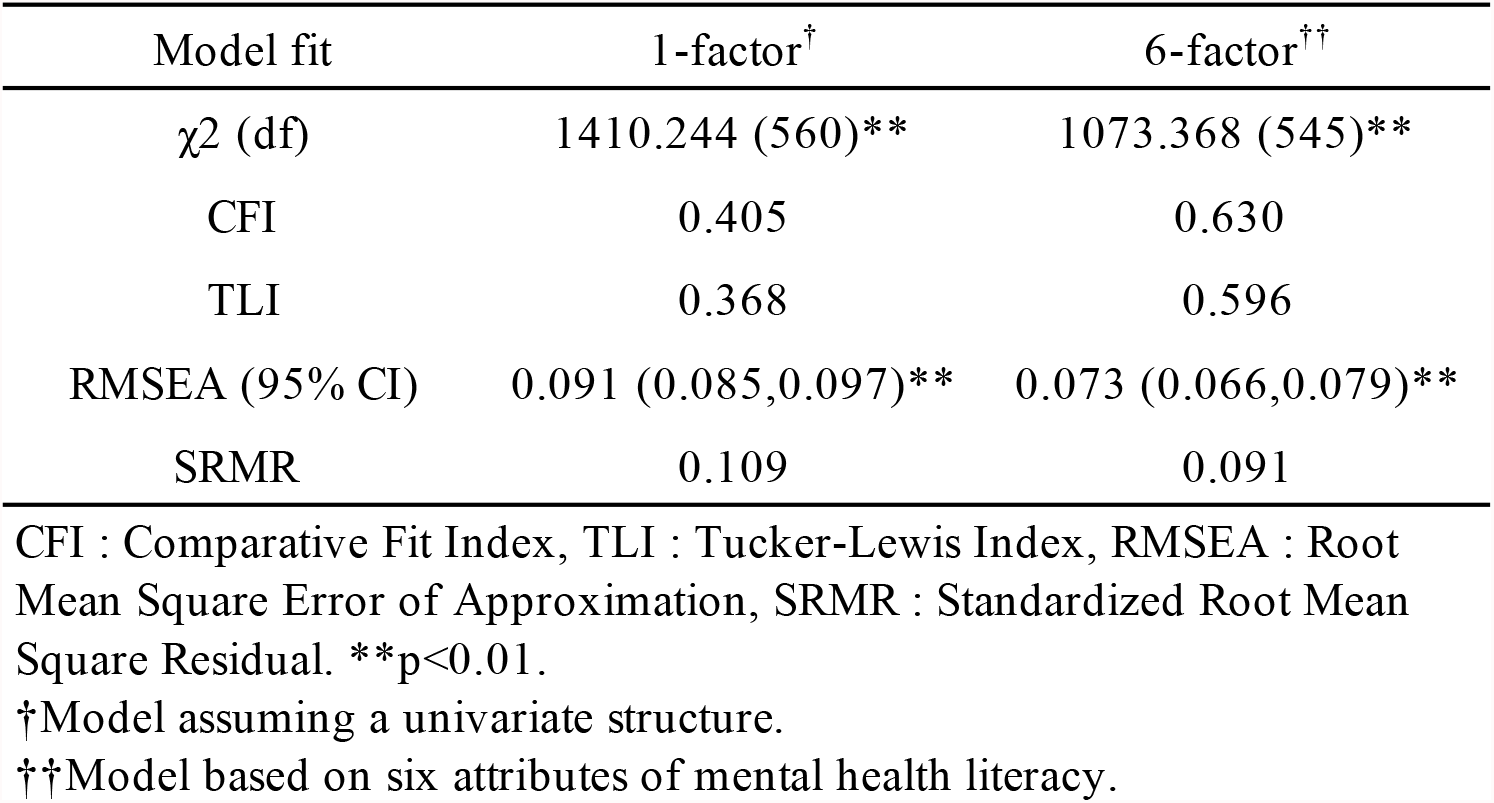
Model fit in confirmatory factor analyses

| Model fit | 1-factor<sup>†</sup> | 6-factor<sup>††</sup> |
| --- | --- | --- |
| $\chi^2$ (df) | 1410.244 (560)** | 1073.368 (545)** |
| CFI | 0.405 | 0.630 |
| TLI | 0.368 | 0.596 |
| RMSEA (95% CI) | 0.091 (0.085,0.097)** | 0.073 (0.066,0.079)** |
| SRMR | 0.109 | 0.091 |

CFI : Comparative Fit Index, TLI : Tucker-Lewis Index, RMSEA : Root Mean Square Error of Approximation, SRMR : Standardized Root Mean Square Residual. \*\*p<0.01.
<sup>†</sup>Model assuming a univariate structure.
<sup>††</sup>Model based on six attributes of mental health literacy.

**Table 4.**
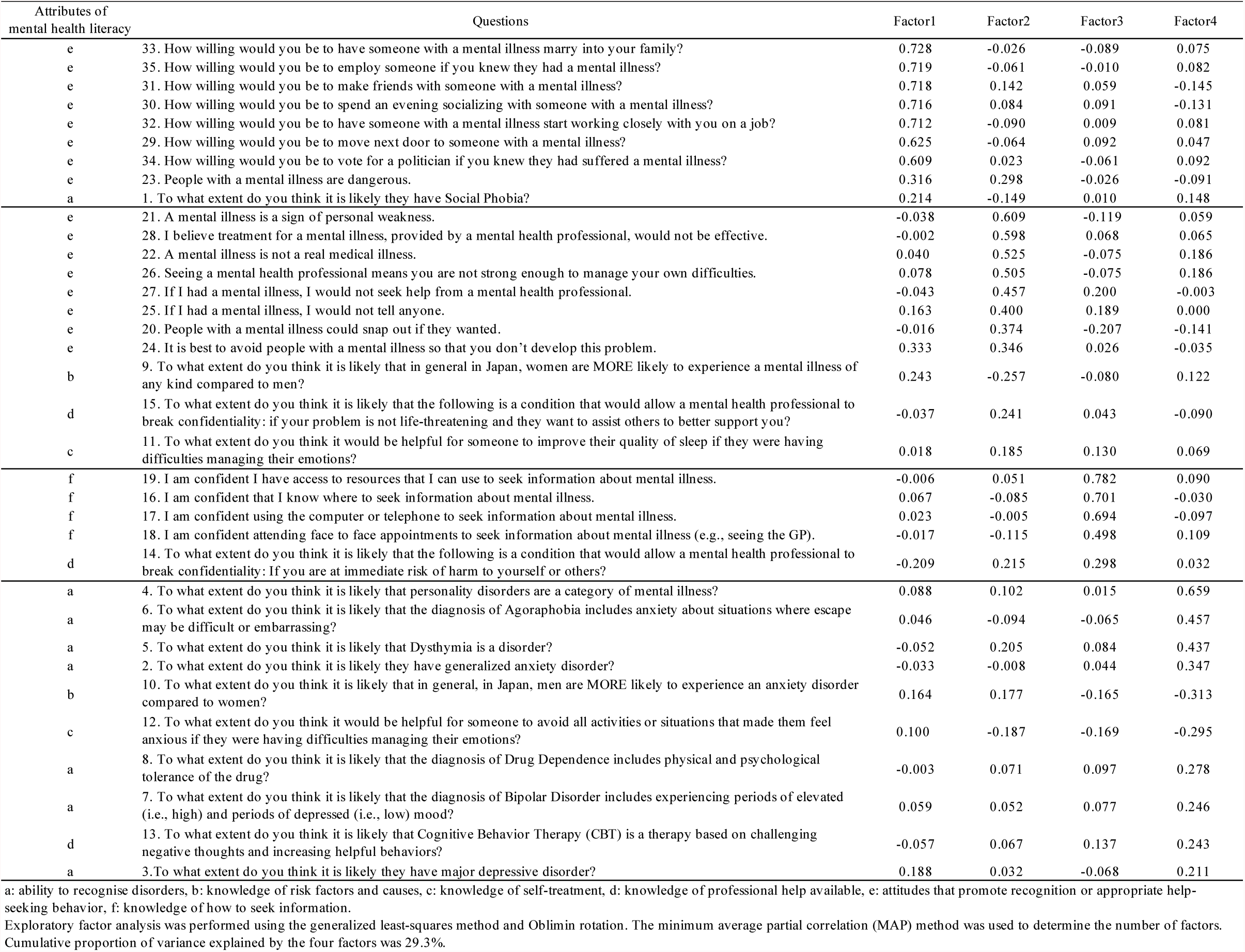
Results of exploratory factor analysis (N = 183)

### Convergent validity

**Table 5** shows Pearson’s correlation coefficients (r) among the MHLS, mental health-related behavior (RIBS), accurate knowledge about mental disorders (MIDUS), intentions to seek help (GHSQ), and psychological distress (K6). The MHLS had a strong positive correlation with mental health-related behavior and moderate positive correlations with accurate knowledge about mental disorders and intentions to seek help. There was no significant correlation with psychological distress.

**Table 5.** Convergent validity of the Japanese version of the MHLS (N = 183)

| Variables | Pearson's correlation with MHLS |
| --- | --- |
| Mental health-related behavior (RIBS) | 0.62** |
| Past or present contact with people with mental health problems | 0.16* |
| Future behavioral intentions when in contact with someone with mental health | 0.65** |
| Accurate knowledge about mental disorders (MIDUS)<sup>†</sup> | -0.31** |
| Treatability of illness<sup>†</sup> | -0.32** |
| Efficacy of medication<sup>†</sup> | -0.13 |
| Social recognition of illness<sup>†</sup> | -0.37** |
| Intentions to seek help (GHSQ) | 0.32** |
| Psychological distress (K6) | 0.14 |

RIBS: Reported and Intended Behaviour Scale, MIDUS: Mental Illness and Disorder Understanding Scale, GHSQ: General Help-Seeking Questionnaire. \*p < 0.05, \*\*p < 0.01.
<sup>†</sup>Lower total scores indicate better understanding, so the correlation is negative.

## Discussion

This study evaluated the reliability and validity of the Japanese version of the MHLS among and graduate students in the medical field. This scale showed good internal consistency, test-retest reliability, and convergent validity. However, the CFA showed that the factor structure did not fit with a univariate structure previously reported^17)^, and the EFA indicated a four-factor structure.

The internal consistency (Cronbach’s α coefficient) of the Japanese version of the MHLS was 0.76, which was slightly lower than that of the original version of MHLS (α = 0.87)^17)^ and close to that of other language versions of the MHLS (Vietnamese (α = 0.72)^20)^, Persian (α = 0.74)^21)^, and Chinese (α = 0.79)^23)^). One of the reasons for the lower results than the original version is the possibility that the translation procedure was not good enough. Although we followed the guidelines, we cannot completely discard this possibility. It is possible that items did not properly express the concept, and the respondents may have interpreted it differently from what the original version indicated. The ICC was 0.77, which could be interpreted as excellent^41)^, and the test-retest reliability was good. These results suggest that the MHLS can be used for different groups with good internal consistency and stability.

The Japanese version of the MHLS showed a moderately significant positive correlation with intention to seek help (GHSQ), which was consistent with the results of the original paper^17)^. The Japanese version of the MHLS also showed a significant positive correlation with mental health-related behavior (RIBS) and accurate knowledge about mental disorders (MIDUS), consistent with the hypothesis. The results were shown to be of good convergent validity. In addition, there was no significant correlation between the MHLS and the psychological distress (K6). This is consistent with the previous report of no correlation between the MHLS and psychological distress as measured by K10 in the original paper^17)^. The level of psychological distress may not be related to the level of MHL. The association between MHL and psychological distress may involve other factors that cannot be explained by eliminating prejudice and promoting early diagnosis.

The CFA did not support either the one-factor model or the six-factor model. While the original authors proposed the one-factor model, they also mentioned a possible four-factor model behind the MHLS^17)^. Thus it may be reasonable that the CFA did not support the one-factor model. In this study, the EFA yielded a four-factor solution. It is unclear if the observed four-factor solution was the same as one that the original authors found^17)^ because they did not provide detailed results for this model. However, the observed four-factor model is approximately consistent with the composition structure of the MHL attributes included in the MHLS. For example, Factor 3 seems related to knowledge of how to seek information (component f); Factor 4 consists of items for ability to recognize disorders (a). The items of attitudes that promote recognition or appropriate help-seeking behavior (e) were grouped together but split into two factors: attitudes toward people with mental disorders and beliefs about mental disorders. These two attributes may be closely related, but classifiable into these two aspects. While the six-factor structure strictly following the MHLS attributes was not supported, the MHLS seems to consist of several theoretically meaningful factors, which may partly support the structural validity. A similar four-factor structure was reported for the Arabic^22)^ and Chinese^23)^ versions. The four-factor structure may best fit the MHLS. However, there may be other possible explanations for the findings that neither the one-factor nor the six-factor structure was supported. The first is the impact of the different characteristics of the participants. The participants in the original paper were mental health professionals and students in a psychology class^17)^, while the participants in this study were undergraduate and graduate students in the medical field. The characteristics of the participants may have resulted in different factor structures. Second, the translated items may be interpreted by respondents differently from what was intended in the original ones, due to a problem in the translation not well considered in the translation process and/or cultural and other contextual differences of meaning of the items between Australia and Japan. As a difference in cultural backgrounds, previous studies comparing Japanese and Australians reported that Japanese are more likely to have negative attitudes toward mental health^42)^ and consider character weakness as risk factors of mental disorders^43)^. There are also differences in the mental health care system, with Japan emphasizing hospital care and Australia emphasizing community care^44)^. Further research on the factor structure of the MHLS is required, with a larger sample size including diverse subgroups across different countries. The translated items in this study may also need to be further revised considering the cultural background and the current mental health context, e.g., through extensive qualitative studies.

This study has some limitations. First, because the response rate of the baseline survey could not be calculated, there may be selection bias. For instance, participants with low MHL may have been reluctant to participate in the survey. Second, the generalization of the results should be carefully considered because the participants were selected from a specific set of universities (around fifteen).

In conclusion, the Japanese version of the MHLS showed good reliability and validity, while the factor structure may not be same as for the original scale^17)^. This scale may be useful for assessing the degree of MHL among students in the medical field in Japan. Further research is needed to replicate the present findings in other samples of students in the medical field as well as other populations.

## Data Availability

All data produced in the present study are available upon reasonable request to the authors

